# Maternal Antibody Response and Transplacental Transfer Following SARS-CoV-2 Infection or Vaccination in Pregnancy

**DOI:** 10.1101/2022.03.17.22272574

**Authors:** Sebastian Otero, Emily S. Miller, Ashwin Sunderraj, Elisheva D. Shanes, Allison Sakowicz, Jeffery A. Goldstein, Leena B. Mithal

**Affiliations:** Ann & Robert H. Lurie Children’s Hospital of Chicago and Stanley Manne Children’s Research Institute, 225 E Chicago Ave, Box #20, Chicago, IL, USA; Department of Obstetrics and Gynecology, Northwestern University Feinberg School of Medicine, 675 N St Clair St Ste 14-200, Chicago, IL, USA; Department of Pediatrics, Northwestern University Feinberg School of Medicine, Chicago, IL, USA; Department of Pathology, Northwestern University Feinberg School of Medicine, 710 N Fairbanks Ct Olson Pavilion, Ste 2-458, Chicago, IL, USA

**Keywords:** SARS-CoV-2, COVID-19, vaccination, pregnancy, antibody

## Abstract

**Background:** Pregnant persons are at increased risk of severe COVID-19 and adverse obstetric outcomes. Understanding maternal antibody response and transplacental transfer after SARS-CoV-2 infection and COVID-19 vaccination is important to inform public health recommendations.

**Methods:** This prospective observational cohort study included 351 birthing individuals who had SARS-CoV-2 infection or COVID-19 vaccination during pregnancy. IgG and IgM to SARS-CoV-2 S1 receptor binding domain were measured in maternal and cord blood. Antibody levels and transplacental transfer ratios were compared across 1) disease severity for those with SARS-CoV-2 infection and 2) infection versus vaccination.

**Findings:** There were 252 individuals with SARS-CoV-2 infection and 99 who received COVID-19 vaccination during pregnancy. Birthing people with more severe SARS-CoV-2 infection category had higher maternal and cord blood IgG levels (p=0.0001, p=0.0001). Median IgG transfer ratio was 0.87-1.2. Maternal and cord blood IgG were higher after vaccination than infection (p=0.001, p=0.001). Transfer ratio was higher after 90 days in the vaccinated group (p<0.001). Modeling showed higher amplitude and half-life of maternal IgG following vaccination (p<0.0001). There were no significant differences by fetal sex.

**Interpretation:** COVID-19 vaccination in pregnancy leads to higher and longer lasting maternal IgG levels, higher cord blood IgG, and higher transfer ratio after 90 days compared to SARS-CoV-2 infection. Greater infection severity leads to higher maternal and cord blood antibodies. Maternal IgG decreases over time following both vaccination and infection, reinforcing the importance of vaccination, even after infection, and vaccine boosters for pregnant patients.

## Introduction

An increasing body of evidence demonstrates that pregnant persons with SARS-CoV-2 infection are at higher risk of severe COVID-19 including hospitalization, intensive care, mechanical ventilation, and death^1-3^. Furthermore, adverse perinatal outcomes such as increased risk of preterm birth, preeclampsia, and stillbirth have been observed with SARS-CoV-2 infection in pregnancy, particularly in the setting of moderate-severe disease^1 3-6^. COVID-19 vaccines have been granted Emergency Use Authorization by the U.S. Food and Drug Administration, although pregnant persons were excluded from published phase three vaccine trials. COVID-19 vaccines include messenger RNA (mRNA) vaccines BNT162b2 (Pfizer/BioNTech) and mRNA-1273 (Moderna) and viral vector vaccines such as J&J/Janssen. Pregnant individuals have had access to vaccines since their availability, with evolving strength of the recommendation and now urgency to protect pregnant persons from COVID-19 through vaccination^7^. There is growing data and literature demonstrating vaccine safety^8-10^ and vaccine efficacy^11 12^ in the pregnant population, yet vaccine hesitancy remains a problem.

Immunologic response, including antibody production, of pregnant individuals following SARS-CoV-2 infection and vaccination has been studied. SARS-CoV-2 infection in pregnancy generates antibody responses, with increase of antibody over many weeks from infection^13-15^. In nonpregnant adults, disease severity is associated with the quantity of circulating antibodies, and antibodies decrease over time after an initial peak^16,17^. However, there is a paucity of pregnancy-specific data on kinetics of antibody titers over time from SARS-CoV-2 infection. Fetal sex may also impact maternal and placental immune response to infection^18^. Additionally, mRNA vaccine immunogenicity in pregnancy has been demonstrated^19 20,21^. There is also a clear need for a second dose for adequate initial vaccine response^19,22,23^. Yet, few studies have addressed the duration of vaccine-induced antibody response in pregnancy or have compared vaccine-induced antibodies to natural infection in pregnancy.

Maternal antibody response serves not only as a marker of the protection for the pregnant person but also correlates with infant passive immunity. Transplacental antibody transfer following infection or vaccine confers protection to the newborn, and vaccination during pregnancy is an important prevention strategy to promote infant health^24^. Transplacental antibody transfer has been shown in the setting of SARS-CoV-2 infection and mRNA vaccination in pregnancy with varying reported transfer ratios (0.3-1.3)^14,25-29^. There have also been concerns of impaired transplacental transfer after SARS-CoV-2 infection^13,30^. With respect to COVID vaccination, two doses of the mRNA vaccine (as opposed to one)^23,27,28^ and vaccination earlier in pregnancy (as opposed to later in the 3^rd^ trimester) are associated with higher transfer ratio, but data outside of third trimester are still limited^31,32^. Given the likely waning of antibody titer and boosters recommended for those at high risk for severe COVID-19, including pregnant individuals, how the timing of infection or vaccination impacts duration of maternal antibody and transplacental antibody transfer is of particular interest.

We investigated maternal anti-spike protein (S1) receptor binding domain (RBD) IgG and IgM of pregnant people and cord blood (herein referred to as infant), at the time of delivery in a large pregnancy cohort with either SARS-CoV-2 infection or mRNA vaccination in pregnancy. We aimed to specifically assess the association between timing/severity of infection and both maternal and infant antibody levels. In addition, we aimed to compare antibody levels between pregnant people who were infected with SARS-CoV-2 versus those with COVID-19 vaccination.

## Methods

### Study design and patient cohort

This is a prospective observational cohort study of pregnant people who delivered at Northwestern Medicine Prentice Women’s Hospital in Chicago, IL, USA (April 2020-July 2021). Individuals who had SARS-CoV-2 infection during their pregnancy or received COVID-19 vaccination were identified via Electronic Medical Record (EMR) review. Maternal SARS-CoV-2 infection during pregnancy was defined by either a SARS-CoV-2 PCR positive laboratory test result or documentation by medical provider of a positive SARS-CoV-2 test result in the EMR. During the study period, all pregnant people were tested for SARS-CoV-2 at the time of admission for delivery, unless they had a prior recent documented infection. Demographic and clinical data, including the presence and absence of specific COVID-19 symptoms, laboratory abnormalities, imaging, clinical course, and treatment were collected by research staff through EMR review. SARS-CoV-2 infection severity was defined according to National Institutes of Health and Society for Maternal-Fetal Medicine criteria as asymptomatic, mild, moderate, severe, and critical illness^33^.

Vaccination during pregnancy and type of vaccine were taken from the EMR that interfaces with Illinois state vaccination record, Illinois Comprehensive Automated Immunization Registry Exchange (I-CARE). Timing of SARS-CoV-2 infection or vaccination was determined by gestational age of SARS-CoV-2 diagnosis or vaccination utilizing the clinically defined estimated due date^34^. For asymptomatic patients, such as those who tested positive on routine PCR screening on admission to Labor and Delivery unit, specific timing of infection could not be accurately determined. Thus, for analyses of related to the timing of infection, these individuals were excluded. The timing of vaccination was based on the gestational age at first dose. Timing from event (infection vs. vaccine) to delivery is termed “latency”. We excluded patients who were both vaccinated and had a SARS-CoV-2 infection in pregnancy. This study was approved by the Institutional Review Board of Northwestern University (reference number STU00212232) with a waiver of informed consent was obtained prior to initiation of this research.

For all analyses, statistical significance was defined as p□<0.05. Corrections were not made for multiple comparisons. Analyses were conducted using STATA/IC version 16.0 (Statacorp), R software, and Python 3.10.1.

### Maternal and infant antibody detection

After birth, maternal blood and umbilical cord blood were retrieved from specimens sent to the clinical laboratory. A maternal blood specimen is sent to the blood bank for all pregnant people upon admission to Labor & Delivery. Umbilical cord blood is sent to the blood bank in the setting of maternal blood type O, Rh negativity, or any positive maternal blood group antibodies. Collected maternal and infant samples were centrifuged; plasma was separated and stored frozen at -80 degrees Celsius. SARS-CoV-2 IgM and IgG were measured in maternal and infant plasma using the Access SARS-CoV-2 IgG and IgM Antibody tests (DXI Platform, Beckman Coulter, Brea CA) in the Northwestern Memorial Hospital CAP/CLIA certified clinical laboratory. The chemiluminescent assay reports quantitative antibodies to the SARS-CoV-2 spike protein S1 RBD in arbitrary units [AU]/mL, with values of ≥ 1 considered positive. Antibody transfer ratio was calculated as infant IgG divided by maternal IgG concentrations.

### Statistical analysis: SARS-CoV-2 severity

Sociodemographic and clinical characteristics were compared between those with SARS-CoV-2 infection across strata of severity. Maternal and infant IgG at the time of delivery as well as the transfer ratio, were compared across severity of infection. Linear regression of maternal and infant IgG levels was performed and locally weighted smoothing (LOESS) smoothing was applied for regression analysis fitting of antibody plots with respect to latency time. As described above, asymptomatic patients were excluded from latency to delivery time analysis because inability to determine the precise timing of infection.

Differences across severity with respect to latency were evaluated using two methods. First, latency from infection was dichotomized at 90 days and median antibody levels were compared across severity subgroups within each of these strata, using Kruskal-Wallis ANOVA. Ninety days was used for clinical relevance (often used as the timeframe reinfection is deemed less likely due to “recent” infection). This analysis was performed separately for maternal IgG and infant IgG. Second, mathematical modeling of maternal antibody durability was conducted using amplitude over time by previously described methods^35^ (code available at https://github.com/jagstein/antibody_kinetics). This approach assumes two antibody secreting populations, one with a long half-life (llp) and the other with a short half-life (slp). In this model, the equation of:

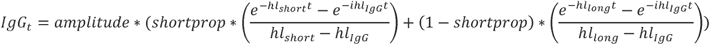

Where IgG_t_ is the IgG level at a latency of t days; amplitude is a measure of intensity; shortprop is the proportion of the IgG-secreting population that is short lived, hl_short_ is the half-life of the short-lived population, hl_IgG_ is the half-life of IgG, and hl_long_ is the half-life of the long-lived population. The proportion of IgG-secreting cells that are long-lived are not directly estimated but are simply the proportion of IgG secreting cells that are not short lived (1 – shortprop). Five-fold cross validation was performed twenty times with shuffling of the IgG analyte (maternal) and categories (mild, moderate, severe, vaccinated). Amplitudes of subcategories were compared using t-tests to report statistically significant differences in antibody kinetics between subgroups.

### Statistical analysis: SARS-CoV-2 infection versus vaccination

A similar approach was used to compare those with SARS-CoV-2 infection versus vaccination. Specifically, sociodemographic and clinical characteristics were compared between those with SARS-CoV-2 infection and those who were vaccinated. Linear regression of maternal and infant IgG levels and LOESS smoothing were performed. In addition, latency from either infection or vaccination was dichotomized at 90 days and median antibody levels (maternal IgG and infant IgG) were compared across these subgroups within each of the latency strata using Mann Whitney U tests. Differences in maternal IgG kinetics were also evaluated using the mathematical modeling approach described above. In patients with multiple inoculations, incorporation of a second stimulus / secretion curve did not improve accuracy, and so was not further pursued.

### Statistical analysis: sex as a biologic variable

Median maternal IgG, infant IgG, and transfer ratios were compared across infant sex using Mann Whitney U tests. These analyses were repeated for those with SARS-CoV-2 infection and those who received COVID-19 vaccination.

## Results

### Study population and characteristics

There were 351 birthing people and 357 infants in the study. Of the birthing people, 252 were diagnosed with SARS-CoV-2 infection and 99 received COVID-19 vaccination during pregnancy. Maternal and neonatal characteristics are reported in Table 1.

**Table 1:** Patient demographics and delivery outcomes.

| Maternal | Total Cohort<br>N = 351 | SARS-CoV-2<br>infection<br>N = 252 | COVID-19<br>vaccine<br>N = 99 | p-value |
| --- | --- | --- | --- | --- |
| Maternal Age<br>mean (std) | 32.5 (5.7) | 31.7 (5.9) | 34.1 (4.9) | <0.001 |
| Race |  |  |  | 0.002 |
| White | 183 (52.1) | 109 (43.3) | 74 (74.8) |  |
| Black | 31 (8.8) | 29 (11.5) | 2 (2.0) |  |
| Asian | 15 (4.3) | 8 (3.2) | 7 (7.1) |  |
| Other | 102 (29.1) | 92 (36.5) | 10 (10.1) |  |
| Unknown | 20 (5.7) | 14 (5.6) | 6 (6.1) |  |
| Latinx | 122 (35.8) | 115 (46.6) | 7 (7.5) | <0.001 |
| Public insurance | 113 (32.2) | 109 (43.3) | 4 (4.0) | <0.001 |
| Multiple Gestations | 5 (1.4) | 4 (1.6) | 1 (1.0) |  |
| Preterm Birth | 37 (10.5) | 32 (12.7) | 5 (5.1) | 0.035 |
| Nulliparous | 129 (35.8) | 105 (41.7) | 24 (24.2) | 0.002 |
| Comorbidities |  |  |  |  |
| Asthma | 40 (11.4) | 35 (13.9) | 5 (5.1) | 0.024 |
| Obesity | 185 (52.7) | 156 (61.9) | 29 (29.3) | <0.001 |
| Tobacco use | 32 (9.1) | 28 (11.1) | 4 (4.0) | 0.040 |
| Chronic Hypertension | 7 (1.99) | 7 (2.78) | 0 | 0.19 |
| Gestational Age at Exposure mean (std) | (8.9) | 29.0 (9.9) | 31.3 (5.3) | 0.62 |
| **Infant** | **N=357** | **SARS-CoV-2 Infection N = 256** | **COVID-19 vaccine N = 101** |  |
| Sex (female) | 181 (50.7) | 134 (52.3) | 47 (46.5) | 0.38 |
| Cord blood available | 298 | 234 | 64 |  |

### Maternal disease severity

Of those with SARS-CoV-2 infection, severity categories were 64 (26%) asymptomatic, 140 (56%) mild disease, 32 (13%) moderate disease, and 12 (5%) severe/critical (Supplementary Table 1). The majority of pregnant people with infections in first and second trimesters were symptomatic: 20 (20/24, 83%) in first trimester and 59 (59/62, 95%) in second trimester. Among 162 people with third trimester infection, 105 (105/162, 65%) were symptomatic. There was no statistically significant difference in median latency between infection and delivery based on severity of SARS-CoV-2 infection (mild 65.5 days (IQR 25-124), moderate 70.5 days (IQR 27-128), severe/critical 60 days (IQR 13-104), p = 0.67). Of pregnant people with SARS-CoV-2 infection, 63% had a positive IgG (> 1 AU/ml) at the time of delivery with median level of 1.67 AU/ml (IQR 0.45-6.24) and subset with symptomatic infection had similar rate of positive 66% with median of 1.71 AU/ml (IQR 0.60-7.79). Infant IgG following maternal infection during pregnancy was positive in 58% with median IgG level 1.28 AU/ml (IQR 0.37-5.86). In subset of pregnant people with symptomatic infection, 63% of infants had positive IgG with median of 1.69 AU/ml (IQR 0.56-7.94).

Pregnant people with more severe SARS-CoV-2 infection were more likely to have higher maternal and infant IgG levels (p = 0.0001, p = 0.0001, Figure 1). Patients with more severe SARS-CoV2 infection were also more likely to have maternal IgG levels > 1 AU/ml (asymptomatic 55% vs mild 61% vs moderate 78% vs severe 92%, p = 0.02), which was also reflected in infant IgG levels (asymptomatic 45% vs mild 60% vs moderate 79% vs severe/critical 80%, p = 0.026). Median IgG transfer ratio ranged from 0.87-1.2 and was not significantly different among severity categories (p = 0.29). The LOESS plots in Figure 2 show the best fit trend of the maternal IgG and infant IgG for patients with symptomatic SARS-CoV-2 infection with respect to latency from infection. Dichotomizing latency from infection by ≤ and > 90 days (Supplementary Table 2), median maternal IgG levels were significantly higher with more severe infection after 90 days of latency (p < 0.001) but did not reach statistical significance at ≤ 90 days (p = 0.085). Similarly, infant IgG was significantly higher when infection was > 90 days prior to delivery (p = 0.004) but not significantly different if latency was ≤ 90 days (p = 0.21). Transfer ratios were not significantly different across severity group (p = 0.78 for ≤ 90 latency group and p = 0.67 for > 90 day latency group). Mathematical modeling demonstrated significantly higher maternal IgG amplitude and longer-half life with greater disease severity category (p < 0.0001 across severity categories and in individual comparisons mild vs. moderate, moderate vs. severe, mild vs. severe). Models are displayed in Supplementary Figure 1A.

**Figure 1.**
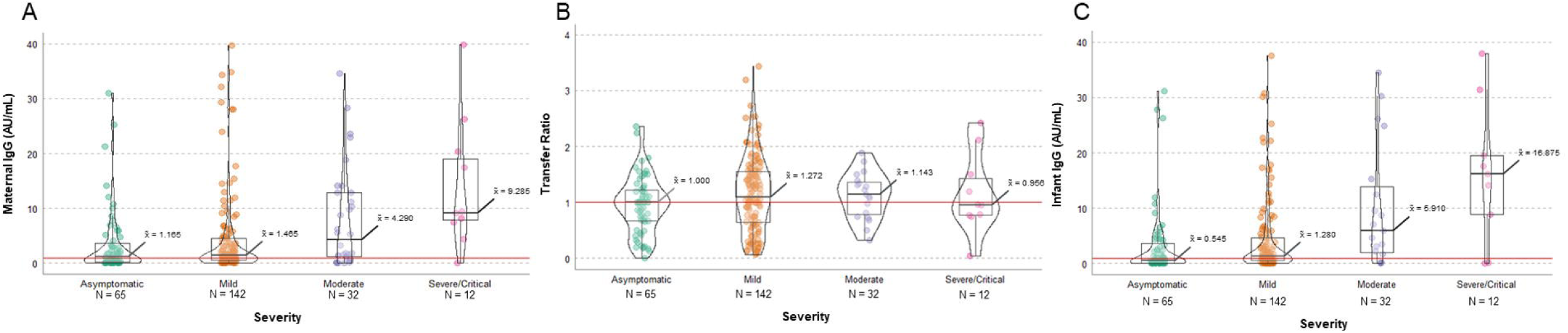
Antibody data by severity of maternal SARS-CoV-2 infection. A) Maternal IgG, B) Transfer ratio (infant IgG/maternal IgG), C) Infant IgG across disease maternal disease severity categories.

**Figure 2.**
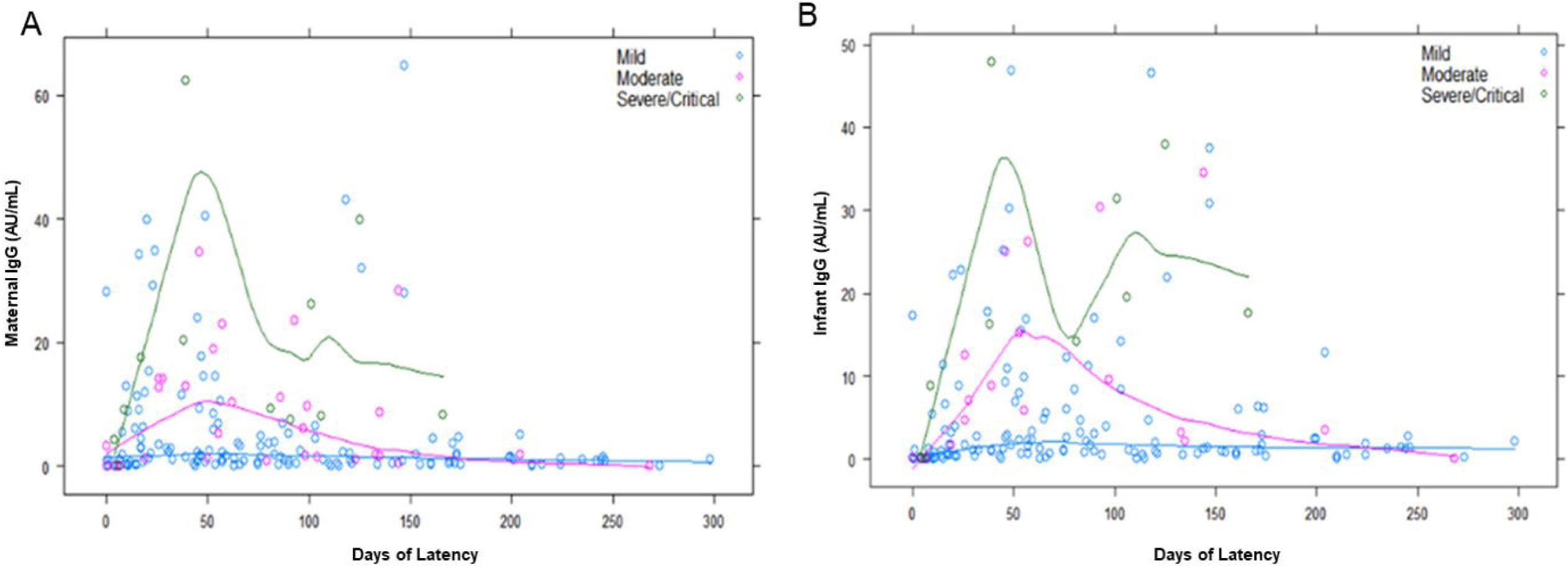
Antibody levels by severity of maternal SARS-CoV-2 infection with respect to latency from SARS-CoV-2 infection to delivery. A) Maternal IgG, B) Infant IgG across disease. Green = severe, pink = moderate, blue = mild.

### SARS-CoV-2 infection and vaccination

Significant differences were noted in maternal age, race/ethnicity, public insurance, parity, asthma, obesity, and tobacco use among the SARS-CoV-2 infection group compared to vaccinated (Table 1). Additionally, there was a higher percentage of preterm births with infection compared to those who were vaccinated (p = 0.035). Of vaccinated patients, 63 (64%) had Pfizer NT162b2, 32 (32%) had Moderna mRNA-1273, and 4 (4%) unknown type. The first dose of vaccination was in second trimester for 17 individuals (17%) and in the third trimester for 82 (83%). Twenty patients (20% of vaccinated group) received only 1 dose of vaccine prior to delivery. The median latency between SARS-CoV-2 infection and delivery was 66 days (IQR 25-124), whereas the median latency between first dose vaccination and delivery was 46 days (IQR 33-71), (p = 0.007).

The vast majority of individuals (92%) had positive maternal IgG following vaccination during pregnancy with median of 16.54 AU/ml (IQR 8.58-28.53). Infant IgG was positive in 89% of infants born to individuals who were vaccinated during pregnancy with median infant IgG 17.38 AU/ml (IQR 7.60-31.26). Overall, maternal IgG levels were higher in the vaccinated group than those with SARS-CoV-2 infection (16.64 vs 1.66, p = 0.001). Infant IgG was also higher in the vaccinated group compared to those with natural infection (17.6 vs 1.29, p = 0.001). Maternal IgM was slightly higher in the vaccinated group (IgM 1.21 vs 0.56, p = 0.001). There was no difference in infant IgM between the two groups (0.15 vs 0.15, p=1.00). Patients who were vaccinated during pregnancy were more likely to have maternal IgG levels > 1 AU/ml compared to those with SARS-CoV-2 infection (92% vs 63%, p < 0.001), a trend which was reflected in infant IgG as well (90% of vaccination group vs. 58% of infection group, p<0.001). Figure 3 demonstrates the distribution of maternal IgG, infant IgG, and transfer ratio following infection and vaccination respectively. LOESS plots in Figure 4 show best fit curves for maternal IgG and infant IgG in the vaccinated and SARS-CoV-2 infection groups by days of latency. Vaccinated individuals had higher maternal IgG and infant IgG levels that appear to be longer lasting, although they appear to downtrend after approximately day 50 and are notably low by approximately 150 days from exposure.

**Figure 3.**
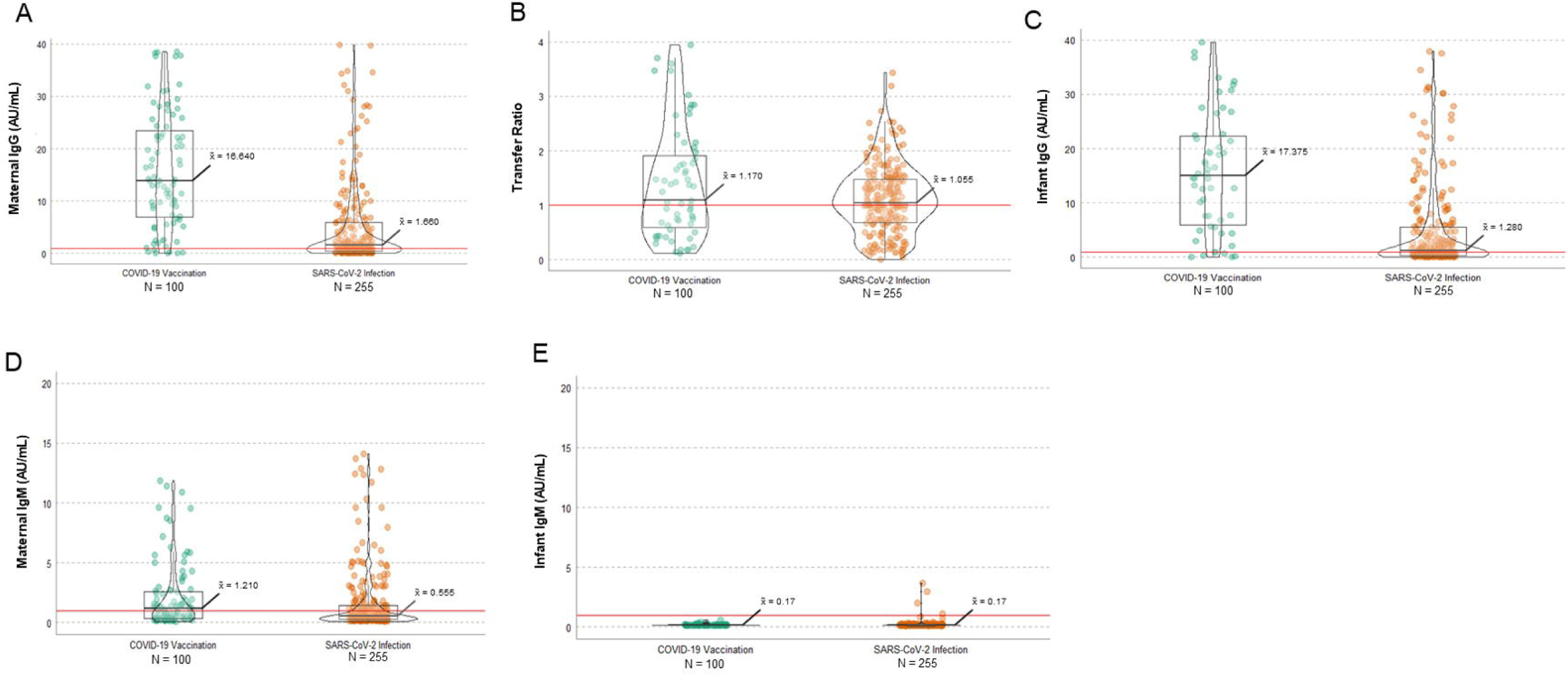
Antibody data in vaccination versus SARS-CoV-2 infection. A) Maternal IgG, B) Transfer ratio (infant IgG/maternal IgG), C) Infant IgG, D) Maternal IgM, E) Infant IgM.

**Figure 4.**
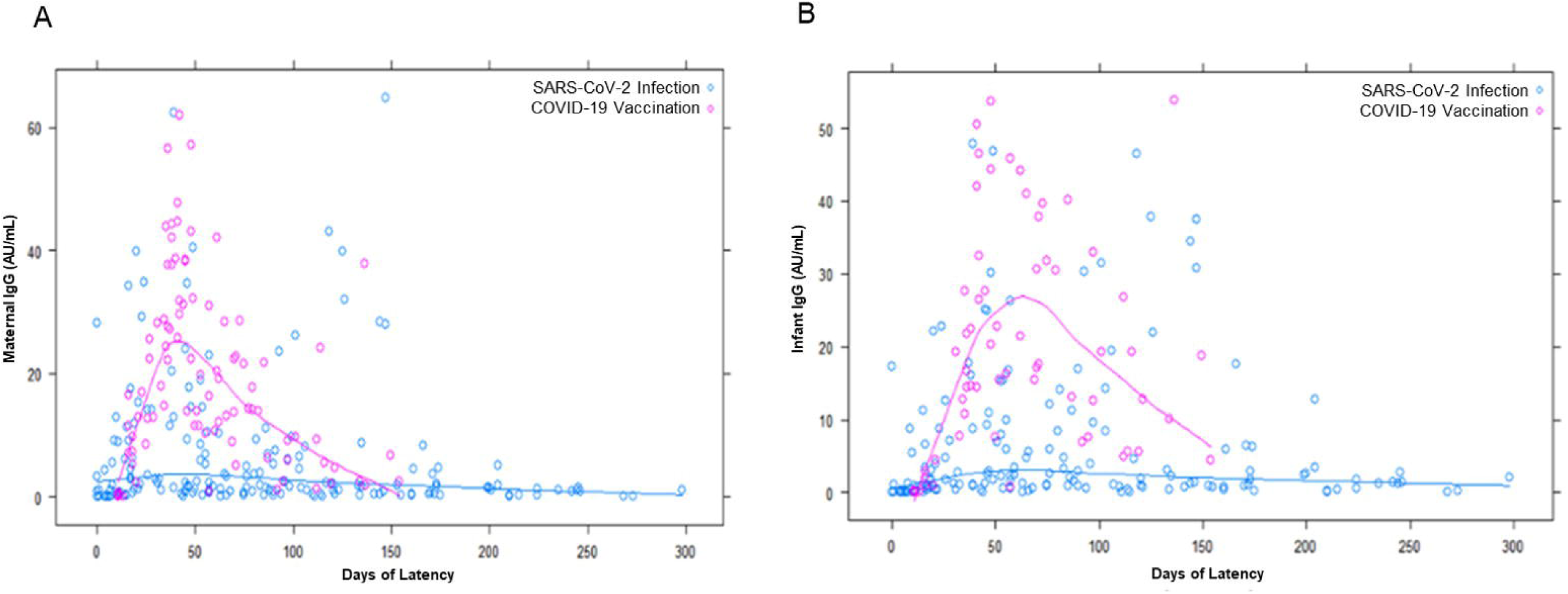
Antibody levels for vaccinated and SARS-CoV-2 infection groups with respect to latency from exposure (first vaccine, infection) to delivery. A) Maternal IgG, B) Infant IgG across disease. pink = vaccination, blue = SARS-coV-2 infection.

The median maternal IgG was significantly higher at delivery in the vaccinated group across latency timeframes (≤ 90 days: 20.40 vs. 2.67, p = 0.001; > 90 days: 5.17 vs. 1.21, p < 0.001). Similarly, infant IgG was significantly higher in vaccinated group (≤ 90 days: 19.79 vs. 1.82, p < 0.001; > 90 days: 12.57 vs. 1.36, p < 0.001). Transfer ratios were not significantly different in the ≤ 90 day timeframe 0.92 vs. 0.94, p = 0.66), but was higher in the > 90 day timeframe (2.84 vs. 1.37, p < 0.001). Mathematical modeling demonstrated significantly higher maternal IgG amplitude and longer-half life in vaccinated people compared to infection (p < 0.0001). Models are displayed in Supplementary Figure 1B.

### Comparison with respect to fetal sex

There were no statistically significant differences by fetal sex in maternal IgG, infant IgG, or transfer ratio in either the vaccinated group, infection group, or pooled analysis of the entire cohort (combing vaccination and infection) (Supplementary Table 3, Supplementary Figure 2).

## Discussion

In this prospective observational cohort study, we addressed maternal and infant antibody levels and transplacental transfer following SARS-CoV-2 infection and vaccination, particularly with respect to latency. Given the concerning body of literature about adverse maternal and obstetric effects of SARS-CoV-2 infection during pregnancy and the protective effects of COVID-19 vaccination, the goal of vaccination is to optimally protect the pregnant person. Thus, it is imperative to understand the durability of protection after SARS-CoV-2 infection and COVID-19 vaccination. Additionally, because infants less than 6 months of age are vulnerable and require a unique vaccination strategy, antibody transfer to the fetus may be an important strategy to optimize protection of young infants from COVID-19.

There is limited data on immune response in relation to disease severity in pregnant people. Prior work by Flannery et al noted that mothers with moderate or critical illness had higher maternal and infant IgG, but the differences were not statistically significant^14^. Our data demonstrates through stratified analysis and modeling that in pregnant people, greater severity of infection is associated with higher and longer lasting maternal antibody levels and higher antibody level in cord blood. It is notable that there was no difference in transplacental antibody transfer with more severe disease, thus the increased levels in umbilical cord blood reflect changes in maternal IgG levels. Furthermore, severity-specific differences in antibody levels are pronounced with longer latency from infection to delivery. Antibody duration has been reported to decrease over time after natural infection, in as little as 3-6 months and particularly after mild infection in nonpregnant individuals^36,37^. In our study, pregnant persons with SARS-CoV-2 infection had a clear rise in IgG with peak approximately 6 weeks from first vaccination (2-3 weeks from second dose) and then a significant fall by 4-5 months, and those with asymptomatic and mild infection yielded lower antibody levels sooner. These data emphasize the need for vaccination as a protective strategy for pregnant people, even after natural SARS-CoV-2 infection during pregnancy.

There is evolving recent literature about the timing and duration of antibody following vaccination in pregnancy. Our study demonstrates that vaccination induces higher and longer lasting antibody levels in pregnant persons and their newborns than SARS-CoV-2 infection, while also avoiding the potential complications of SARS-CoV-2 in pregnancy. The timing of vaccination, duration of response, and transplacental transfer to infant has shown various results across studies. Yang et al found that early third trimester vaccination led to highest anti-spike IgG levels^31^. Kugelman et al reported that second trimester mRNA vaccination led to protective IgG levels at delivery^38^, while Rottenstreich et al reported that IgG levels were lowest after first trimester and highest after 3^rd^ trimester vaccination^32^. Our data, although limited to second and third trimester vaccination, shows clear evidence that vaccination results in superior antibody levels and transfer ratios compared to infection, and that this difference is pronounced with longer latency. Nevertheless, there are decreasing antibody levels over time and, thus this study reinforces the importance of boosting maternal immunity with additional dose if greater than 5 months from primary vaccine series.

In translational work by Bordt et al, mothers carrying male fetuses had lower maternal IgG antibodies and transfer ratio following SARS-CoV-2 infection than pregnancies with female fetuses^18^. The possibility that fetal sex modifies maternal antibody response or placental transfer is intriguing. Our data, however, did not find any statistically significant differences in maternal IgG level or transfer ratio according to fetal sex after SARS-CoV-2 infection or vaccination. Given the hypothesis that maternal proinflammatory response in the setting of acute infection impacts humoral and placental immune response, the timing of infection may be important in fetal sex-specific differences. Bordt and colleagues study focused on acute infection in the third trimester. Our stratified analysis of maternal and infant antibodies limited to third trimester SARS-CoV-2 infection still did not recapitulate the fetal sex-specific differences previously reported. Further dedicated work is needed in this area.

The strengths of this study include 1) the large cohort of samples from pregnant people and umbilical cords in individuals with vaccination or infection during pregnancy, 2) accurate detailed clinical information about spectrum of infection severity and timing, and 3) consistent platform for anti-RBD IgG across all samples in the same laboratory. Additionally, the mathematical modeling approach to maternal antibody kinetics adds to the statistical validity of severity and vaccine vs. natural infection comparisons. Limitations include the single timepoint of antibody measurements at delivery per patient and the single center study. Circulating anti-S1 RBD antibody is also only one measure of immune protection and does not account for aspects of immunologic memory. Although we did not specifically measure neutralizing antibody, anti-S IgG correlates closely with neutralizing activity patterns^16,38^. In addition, given the timeframe of the cohort, the effects of 3^rd^ dose booster vaccination during pregnancy are not assessed.

In conclusion, COVID-19 vaccination in pregnancy leads to higher and longer lasting maternal IgG and higher infant IgG levels than natural SARS-CoV-2 infection. Vaccination results in higher transplacental antibody transfer ratios than infection after longer latency. Severity of infection is associated with higher and longer lasting antibodies. Maternal IgG antibody levels decrease over time in pregnant patients following both vaccination and natural infection, reinforcing the importance of vaccination even after infection and booster doses if 5 months have elapsed from initial series, to optimize protection of pregnant individuals and their infants.

## Data Availability

All data produced in the present study are available upon reasonable request to the authors.

## Acknowledgements

The authors acknowledge the staff of the Northwestern Prentice Women’s Hospital Obstetric COVID Unit, the Northwestern Memorial Hospital Blood Bank and the Northwestern Memorial Hospital Chemistry Lab, for sample processing and antibody assays making this study possible. We also acknowledge contributions of students and research staff who participated in data collection: Antonia Willnow, Rebecca Ebbott, Raveena Aggarwal, Hooman Azad, Chiedza Mupanomunda, Alexandra Isaia, and Allaa Fadl-Alla.

## References

1. Woodworth KR, Olsen EO, Neelam V, et al. Birth and Infant Outcomes Following Laboratory-Confirmed SARS-CoV-2 Infection in Pregnancy - SET-NET, 16 Jurisdictions, March 29-October 14, 2020. MMWR Morb Mortal Wkly Rep 2020; 69(44): 1635–40.

2. Zambrano LD, Ellington S, Strid P, et al. Update: Characteristics of Symptomatic Women of Reproductive Age with Laboratory-Confirmed SARS-CoV-2 Infection by Pregnancy Status - United States, January 22-October 3, 2020. MMWR Morb Mortal Wkly Rep 2020; 69(44): 1641–7.

3. Metz TD, Clifton RG, Hughes BL, et al. Association of SARS-CoV-2 Infection With Serious Maternal Morbidity and Mortality From Obstetric Complications. JAMA 2022; 327(8): 748–59.

4. Karasek D, Baer RJ, McLemore MR, et al. The association of COVID-19 infection in pregnancy with preterm birth: A retrospective cohort study in California. Lancet Reg Health Am 2021; 2: 100027.

5. Lai J, Romero R, Tarca AL, et al. SARS-CoV-2 and the subsequent development of preeclampsia and preterm birth: evidence of a dose-response relationship supporting causality. Am J Obstet Gynecol 2021; 225(6): 689–93 e1.

6. DeSisto CL, Wallace B, Simeone RM, et al. Risk for Stillbirth Among Women With and Without COVID-19 at Delivery Hospitalization - United States, March 2020-September 2021. MMWR Morb Mortal Wkly Rep 2021; 70(47): 1640–5.

7. Centers for Disease Control and Prevention. COVID-19 Vaccines While Pregnant or Breastfeeding.

8. Shimabukuro TT, Kim SY, Myers TR, et al. Preliminary Findings of mRNA Covid-19 Vaccine Safety in Pregnant Persons. N Engl J Med 2021; 384(24): 2273–82.

9. Kharbanda EO, Haapala J, DeSilva M, et al. Spontaneous Abortion Following COVID-19 Vaccination During Pregnancy. JAMA 2021; 326(16): 1629–31.

10. Lipkind HS, Vazquez-Benitez G, DeSilva M, et al. Receipt of COVID-19 Vaccine During Pregnancy and Preterm or Small-for-Gestational-Age at Birth - Eight Integrated Health Care Organizations, United States, December 15, 2020-July 22, 2021. MMWR Morb Mortal Wkly Rep 2022; 71(1): 26–30.

11. Goldshtein I, Nevo D, Steinberg DM, et al. Association Between BNT162b2 Vaccination and Incidence of SARS-CoV-2 Infection in Pregnant Women. JAMA 2021; 326(8): 728–35.

12. Morgan JA, Biggio JR, Jr., Martin JK, et al. Maternal Outcomes After Severe Acute Respiratory Syndrome Coronavirus 2 (SARS-CoV-2) Infection in Vaccinated Compared With Unvaccinated Pregnant Patients. Obstet Gynecol 2022; 139(1): 107–9.

13. Edlow AG, Li JZ, Collier AY, et al. Assessment of Maternal and Neonatal SARS-CoV-2 Viral Load, Transplacental Antibody Transfer, and Placental Pathology in Pregnancies During the COVID-19 Pandemic. JAMA Netw Open 2020; 3(12): e2030455.

14. Flannery DD, Gouma S, Dhudasia MB, et al. Assessment of Maternal and Neonatal Cord Blood SARS-CoV-2 Antibodies and Placental Transfer Ratios. JAMA Pediatr 2021; 175(6): 594–600.

15. Cosma S, Carosso AR, Corcione S, et al. Longitudinal analysis of antibody response following SARS-CoV-2 infection in pregnancy: From the first trimester to delivery. J Reprod Immunol 2021; 144: 103285.

16. Legros V, Denolly S, Vogrig M, et al. A longitudinal study of SARS-CoV-2-infected patients reveals a high correlation between neutralizing antibodies and COVID-19 severity. Cell Mol Immunol 2021; 18(2): 318–27.

17. Wang K, Long QX, Deng HJ, et al. Longitudinal Dynamics of the Neutralizing Antibody Response to Severe Acute Respiratory Syndrome Coronavirus 2 (SARS-CoV-2) Infection. Clin Infect Dis 2021; 73(3): e531–e9.

18. Bordt EA, Shook LL, Atyeo C, et al. Maternal SARS-CoV-2 infection elicits sexually dimorphic placental immune responses. Sci Transl Med 2021; 13(617): eabi7428.

19. Beharier O, Plitman Mayo R, Raz T, et al. Efficient maternal to neonatal transfer of antibodies against SARS-CoV-2 and BNT162b2 mRNA COVID-19 vaccine. J Clin Invest 2021; 131(19).

20. Collier AY, McMahan K, Yu J, et al. Immunogenicity of COVID-19 mRNA Vaccines in Pregnant and Lactating Women. JAMA 2021; 325(23): 2370–80.

21. Prabhu M, Murphy EA, Sukhu AC, et al. Antibody Response to Coronavirus Disease 2019 (COVID-19) Messenger RNA Vaccination in Pregnant Women and Transplacental Passage Into Cord Blood. Obstet Gynecol 2021; 138(2): 278–80.

22. Gray KJ, Bordt EA, Atyeo C, et al. Coronavirus disease 2019 vaccine response in pregnant and lactating women: a cohort study. Am J Obstet Gynecol 2021; 225(3): 303 e1–e17.

23. Atyeo C, DeRiso EA, Davis C, et al. COVID-19 mRNA vaccines drive differential antibody Fc-functional profiles in pregnant, lactating, and nonpregnant women. Sci Transl Med 2021; 13(617): eabi8631.

24. Munoz FM, Jamieson DJ. Maternal Immunization. Obstet Gynecol 2019; 133(4): 739–53.

25. Rottenstreich A, Zarbiv G, Oiknine-Djian E, Zigron R, Wolf DG, Porat S. Efficient Maternofetal Transplacental Transfer of Anti-Severe Acute Respiratory Syndrome Coronavirus 2 (SARS-CoV-2) Spike Antibodies After Antenatal SARS-CoV-2 BNT162b2 Messenger RNA Vaccination. Clin Infect Dis 2021; 73(10): 1909–12.

26. Gray KJ, Bordt EA, Atyeo C, et al. COVID-19 vaccine response in pregnant and lactating women: a cohort study. medRxiv 2021.

27. Mithal LB, Otero S, Shanes ED, Goldstein JA, Miller ES. Cord blood antibodies following maternal coronavirus disease 2019 vaccination during pregnancy. Am J Obstet Gynecol 2021; 225(2): 192–4.

28. Beharier O, Plitman Mayo R, Raz T, et al. Efficient maternal to neonatal transfer of antibodies against SARS-CoV-2 and BNT162b2 mRNA COVID-19 vaccine. J Clin Invest 2021; 131(13).

29. Song D, Prahl M, Gaw SL, et al. Passive and active immunity in infants born to mothers with SARS-CoV-2 infection during pregnancy: prospective cohort study. BMJ Open 2021; 11(7): e053036.

30. Atyeo C, Pullen KM, Bordt EA, et al. Compromised SARS-CoV-2-specific placental antibody transfer. Cell 2021; 184(3): 628–42 e10.

31. Yang YJ, Murphy EA, Singh S, et al. Association of Gestational Age at Coronavirus Disease 2019 (COVID-19) Vaccination, History of Severe Acute Respiratory Syndrome Coronavirus 2 (SARS-CoV-2) Infection, and a Vaccine Booster Dose With Maternal and Umbilical Cord Antibody Levels at Delivery. Obstet Gynecol 2022; 139(3): 373–80.

32. Rottenstreich A, Zarbiv G, Oiknine-Djian E, et al. The effect of gestational age at BNT162b2 mRNA vaccination on maternal and neonatal SARS-CoV-2 antibody levels. Clin Infect Dis 2022.

33. National Institutes of Health. COVID-19 Treatment Guidlines: Clinical Spectrum of SARS-CoV-2 Infection. 2021.

34. Committee Opinion No 700: Methods for Estimating the Due Date. Obstet Gynecol 2017; 129(5): e150–e4.

35. White M, Idoko O, Sow S, et al. Antibody kinetics following vaccination with MenAfriVac: an analysis of serological data from randomised trials. Lancet Infect Dis 2019; 19(3): 327–36.

36. Long QX, Tang XJ, Shi QL, et al. Clinical and immunological assessment of asymptomatic SARS-CoV-2 infections. Nat Med 2020; 26(8): 1200–4.

37. Choe PG, Kim KH, Kang CK, et al. Antibody Responses 8 Months after Asymptomatic or Mild SARS-CoV-2 Infection. Emerg Infect Dis 2021; 27(3): 928–31.

38. Kugelman N, Nahshon C, Shaked-Mishan P, et al. Maternal and Neonatal SARS-CoV-2 Immunoglobulin G Antibody Levels at Delivery After Receipt of the BNT162b2 Messenger RNA COVID-19 Vaccine During the Second Trimester of Pregnancy. JAMA Pediatr 2021.

