## Supplemental for "Maternal Antibody Response and Transplacental Transfer Following SARS-CoV-2 Infection or Vaccination in Pregnancy"

### Supplementary Material

#### Supplementary Table 1: Demographic data and delivery outcomes by SARS-CoV-2 infection severity

| **Maternal** | **Asymptomatic N = 64** | **Mild N = 140** | **Moderate N = 32** | **Severe/Critical N = 12** | **p-value** |
|---|---|---|---|---|---|
| Maternal Age mean (std) | 29.6 (6.2) | 32.1 (5.6) | 33.3 (5.9) | 33.5 (5.7) | 0.43 |
| Race | | | | | 0.096 |
| White | 23 (35.9) | 59 (42.1) | 18 (56.3) | 5 (41.7) | |
| Black | 16 (25.0) | 11 (7.9) | 1 (3.1) | 1 (8.3) | |
| Asian | 2 (3.1) | 6 (4.3) | 0 | 0 | |
| Other | 21 (32.8) | 53 (37.9) | 12 (37.5) | 6 (50.0) | |
| Unknown | 2 (3.1) | 11 (7.9) | 1 (3.1) | 0 | |
| Latinx | 27 (42.2) | 61 (45.2) | 20 (62.5) | 7 (58.3) | 0.22 |
| Public insurance | 38 (59.4) | 52 (37.4) | 14 (43.8) | 5 (41.7) | 0.030 |
| Multiple Gestations | 1 (1.6) | 2 (1.4) | 0 | 1 (8.3) | 0.31 |
| Preterm Birth | 11 (17.2) | 13 (9.3) | 5 (15.6) | 3 (25.0) | 0.16 |
| Nulliparous | 27 (42.2) | 59 (42.1) | 12 (37.5) | 5 (41.7) | 0.98 |
| Comorbidities | | | | | |
| Asthma | 11 (17.2) | 15 (10.7) | 6 (18.8) | 3 (25.0) | 0.23 |
| Obesity | 34 (53.1) | 82 (58.5) | 26 (81.3) | 11 (91.7) | 0.005 |
| Tobacco use | 2 (3.1) | 17 (12.1) | 6 (18.8) | 2 (16.7) | 0.04 |
| Chronic Hypertension | 0 | 4 (2.9) | 2 (6.3) | 1 (8.3) | 0.10 |
| Gestational age at SARS-CoV-2 diagnosis Mean (std) | 35.0 (8.5) | 26.8 (9.7) | 26.9 (9.2) | 28.4 (7.1) | <0.001 |
| **Infant** | **Asymptomatic N = 65** | **Mild N = 142** | **Moderate N = 32** | **Severe/Critical N = 13** | |
| Sex (female) | 34 (52.3) | 71 (50.0) | 18 (56.3) | 9 (69.2) | 0.59 |
| Cord blood available | 64 | 138 | 19 | 10 | |
| Latency time (weeks) med (IQR) | N/A | 9.36 (3.57-17.64) | 10.07(3.86-18.29) | 8.57 (1.86-14.79) | |

^Severity according to NIH guidelines[31]

#### Supplementary Table 2: IgG and transfer ratio comparisons based on severity and latency

| | Latency ≤ 90 days | | | | Latency > 90 days | | | |
|---|---|---|---|---|---|---|---|---|
| | Mild | Moderate | Severe/Critical | p-value | Mild | Moderate | Severe/Critical | p-value |
| Maternal IgG | 1.92 (0.76-6.33) | 7.80 (0.86-14.10) | 20.40 (11.44-31.01) | 0.085 | 0.98 (0.40-2.15) | 1.84 (1.48-8.70) | 8.28 (8.07-26.26) | <0.001 |
| Transfer Ratio | 0.99 (0.35-1.50) | 0.89 (0.70-1.12) | 0.78 (0.75-0.96) | 0.78 | 1.35 (1.04-1.70) | 1.39 (1.29-1.73) | 1.70 (1.07-2.27) | 0.67 |
| Infant IgG | 1.64 (0.46-5.73) | 6.48 (0.90-13.89) | 11.48 (0.19-16.18) | 0.21 | 1.26 (0.65-3.43) | 3.48 (2.20-30.30) | 25.48 (18.55-34.66) | 0.0038 |

Kruskal-Wallis ANOVA

**Supplementary Table 3: IgG and transfer ratio comparisons based on fetal sex**

| | All | | | SARS-CoV-2 Infection | | | COVID-19 Vaccination | | |
|---|---|---|---|---|---|---|---|---|---|
| Med (IQR) | Female | Male | p-value | Female | Male | p-value | Female | Male | p-value |
| Maternal IgG | 3.06 (0.99 - 12.79) | 3.67 (0.54-14.66) | 0.87 | 1.79 (0.62-6.03) | 1.21 (0.39-8.12) | 0.49 | 14.11 (5.83-31.86) | 17.97 (9.40-26.47) | 0.98 |
| Infant IgG | 2.63 (0.54-15-26) | 2.24 (0.46-11.32) | 0.41 | 1.73 (0.37-6.63) | 1.10 (0.41-4.67) | 0.27 | 22.05 (7.56-36.77) | 16.22 (7.76-22.47) | 0.33 |
| Transfer | 1.26 (0.68-1.75) | 1.00 (0.49-1.56) | 0.14 | 1.18 (0.65-1.67) | 1.00 (0.45-1.42) | 0.18 | 1.42 (0.94-2.16) | 0.92 (0.51-2.06) | 0.41 |

Wilcoxon rank sum

**Supplementary Figure 1: Mathematical models of maternal IgG level**

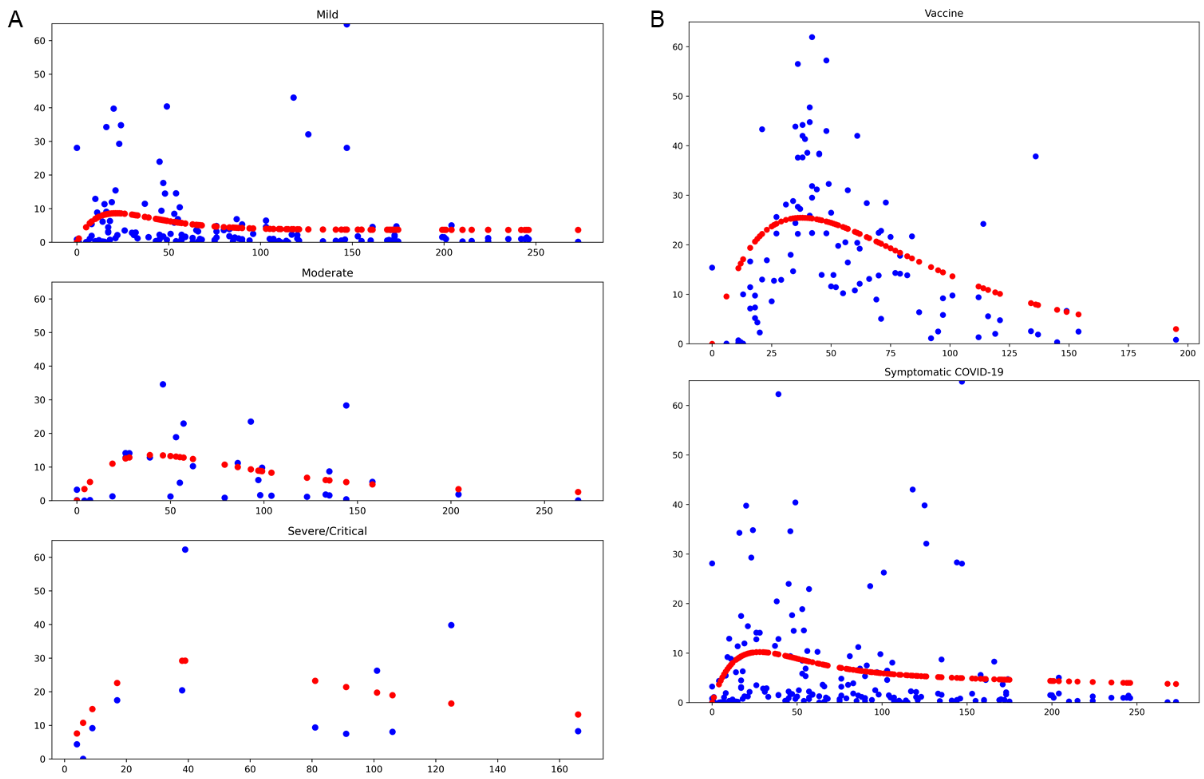


Mathematical models of maternal IgG level. A) across severity categories, B) vaccine vs. symptomatic SARS-CoV-2 infection. Blue = individual data points, red = kinetics model

**Supplementary Figure 2: Antibody data by fetal sex.**

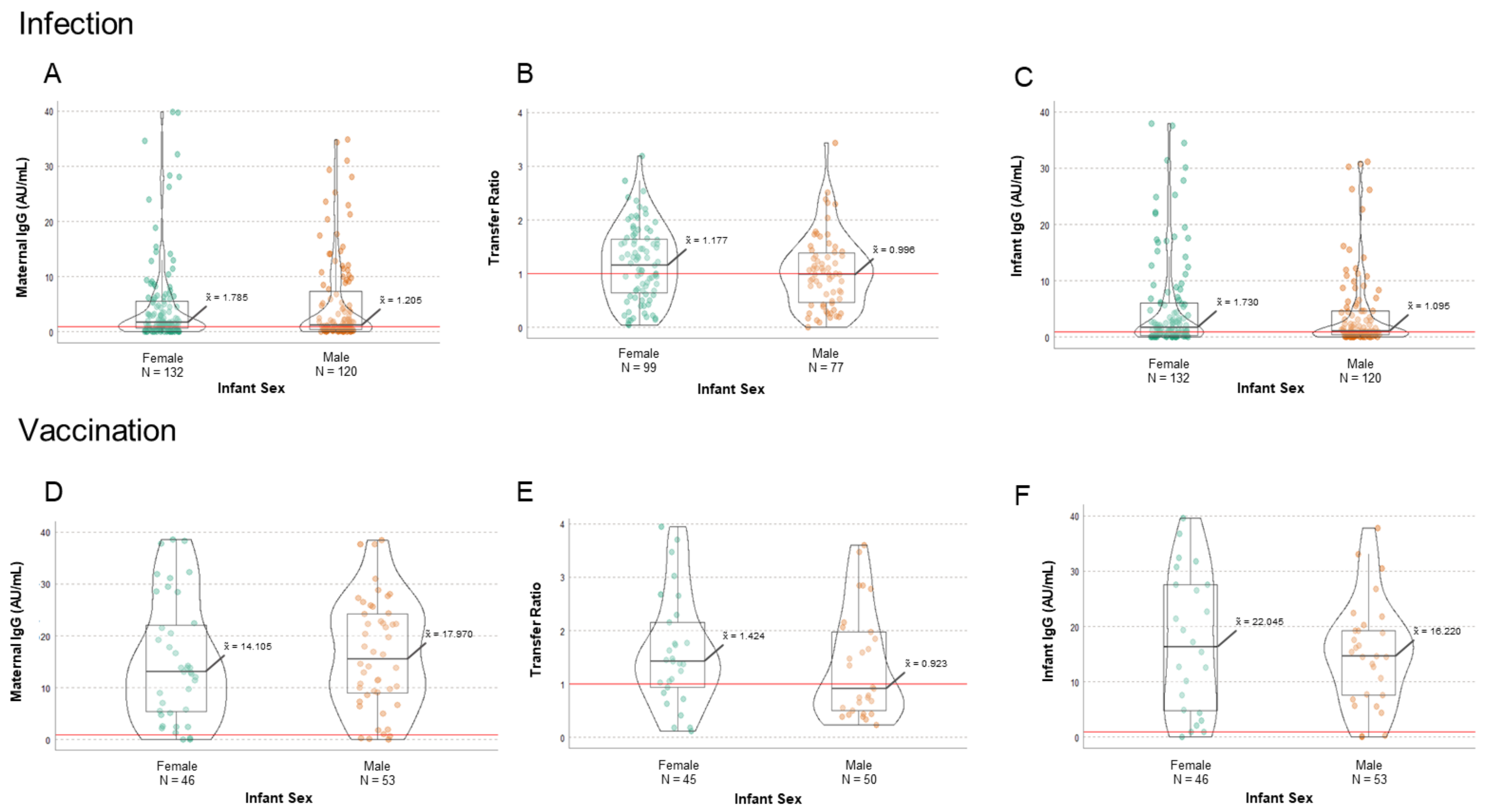


Antibody data by fetal sex. SARS-CoV-2 infection. A) Maternal IgG, B) Transfer ratio (infant IgG/maternal IgG), C) Infant IgG; Vaccination D) Maternal IgG, E) Transfer ratio, F) Infant IgG by fetal sex (female vs. male).
